# Semi-automated quantification of ^18^F-FDG PET-CT in pulmonary TB contacts and its association with prospective outcomes

**DOI:** 10.1101/2025.08.04.25331303

**Authors:** Jee Whang Kim, Anver Kamil, Joanne Lee, Ismail Novsarka, Pranabashis Haldar

## Abstract

Positron Emission Tomography-Computed Tomography (PET-CT) has shown potential as a research tool to characterise heterogeneity in tuberculosis (TB) infection. Prospective utilisation of this technology will require standardisation of imaging protocols, interpretation and analysis of PET-CT images to improve generalisability and data assimilation between discrete cohorts. We evaluated a semi-automated approach using open-source software to minimise inter-operator variability of imaging interpretation and use reference organ normalisation to lower variability in physiological uptake.

We quantified ^18^F-Fluorodeoxyglucose (FDG) uptake in intrathoracic lymph nodes (ITLNs) of eleven QuantiFERON-TB Gold-Plus positive TB contacts that enrolled on a prospective observational study, including eight contacts with serial PET-CT scans. The imaging was analysed using 3D Slicer, an open-source medical imaging platform designed for biomedical and clinical research, that provides semi-automated segmentation and automated liver quantification. The extracted data included ITLN maximum standardised uptake value (SUVmax), mean standardised uptake value (SUVmean), metabolic volume (MV), and total lesion glycolysis (TLG), and liver SUVmean. We compared its performance to that of clinical software, both with and without standardising to liver uptake (as a physiological reference), and assessed operator variability. Then, we evaluated the association between multiple quantitative PET metrics and prospective outcomes in pulmonary TB contacts.

We report strong agreement between open-source and clinical image evaluation platforms. We find semi-automated PET quantification can lower inter-operator variability, and standardising to reference organs increases the sensitivity of imaging analysis. Specifically, we report preliminary findings that increasing metabolic activity on serial PET-CT in early infection is associated with detectable Mtb at the anatomical site of FDG uptake, consistent with a progressive phenotype of infection.

## INTRODUCTION

Tuberculosis (TB) is an infectious disease caused by bacteria of the *Mycobacterium tuberculosis* complex (MTBC), with *Mycobacterium tuberculosis* (Mtb) being the most common pathogen. TB disease continues to pose a significant burden on global health, affecting approximately 10.8 million people in 2023^1^. In the same year, it was responsible for an estimated 1.25 million deaths globally, including 161,000 deaths among people living with HIV^1^.

TB infection is acquired when susceptible hosts inhale droplets containing MTBC from an infected person. A small proportion of those infected develop active TB disease, known as primary progressive tuberculosis. However, in the majority of cases, the infection is successfully controlled by the adaptive immune system, a state referred to as TB infection, or latent tuberculosis infection (LTBI)^2^. TB infection encompasses a spectrum of infection states with varying risk of progression to active disease. Preventing the progression of TB infection to active disease is crucial to meeting the End TB Strategy goal for TB eradication^3,4^. However, the immunological mechanisms that determine the trajectory of TB infection remain poorly understood.

Positron emission tomography-computed tomography (PET-CT) has emerged as a promising research tool to reveal underlying heterogeneity in TB infection states^5-7^. We have previously described PET-CT based phenotype of pre-clinical infection and suggested that incorporating metabolic and structural characterisation could offer clinically relevant stratification^8^. However, there is significant heterogeneity in the analysis of PET-CT imaging in TB infection research, limiting the comparison^9^. Importantly, changes in metabolic activity observed on serial imaging are likely to be smaller in asymptomatic individuals with TB infection, necessitating the need for approaches that accurately assess global metabolic burden.

Normalisation using reference organs can reduce variability in ^18^F-Fluorodeoxyglucose (FDG) uptake in interval scans^10^. However, manual measurement of FDG uptake in the reference organ can also introduce variability in the threshold^11^. An automated technique to determine the volume of interest (VOI) in the reference organ has been proposed to minimise the inter-operator variability^12^. In addition, semi-automated segmentation of target lesions can further reduce variability in measurement^13^.

PET-derived metabolic activity in human TB research is often assessed using the maximum standardised uptake value (SUVmax). Although SUVmax is a widely used marker of disease activity and treatment response, it can overestimate metabolic activity due to image noise^14^, and a two-dimensional region of interest (ROI) may not capture the pixel with the highest value^15^. In this context, volumetric parameters such as metabolic volume (MV) and total lesion glycolysis (TLG) have been reported to be prognostic markers for treatment outcome in active TB^16^. Specifically, TLG considers both mean standardised uptake value (SUVmean) and metabolic volume, thereby offering a composite measure of metabolic activity within a defined volume of interest (VOI).

In this study, we present methodologies for employing an open-source research imaging platform^17^, which provides semi-automated lesion segmentation and automated liver quantification. We apply this approach to the analysis of serial PET-CT scans from asymptomatic, QuantiFERON-TB Gold Plus (QFT-Plus) positive pulmonary TB contacts enrolled in a prospective observational study^8^. The study design has been described previously^18^. We compare the performance of this research platform to that of clinical software, assess inter-operator variability, and explore the association between PET parameters and prospective outcomes.

## PROTOCOL

### 1. PET-CT procedures

1. **Patient Identification** Confirm the patient’s identity using a 3-point ID check (full name, date of birth, and address) against the referral and imaging request.
2. **Patient Safety Questionnaire** Review and complete the patient safety screening questionnaire, ensuring all relevant medical history, allergies, pregnancy status, and contraindications are addressed.
3. **Clinical History Verification** Confirm and document the patient’s clinical history to ensure alignment with the referral and imaging requirements.
4. **Procedure Explanation** Clearly explain the procedure, including:
  - Purpose of the PET scan
  - Injection of a radioactive tracer
  - Associated radiation risks and safety precautions
5. **Informed Consent** Ensure the patient understands the procedure and risks, then obtain written informed consent.
6. **IV Cannulation** Insert an intravenous (IV) catheter, preferably into the median cubital, cephalic, or basilic vein at the antecubital fossa using aseptic technique.
7. **Confirm Vein Patency** Flush the catheter with 10 mL of sterile saline to confirm vein patency. If unsure, consult a colleague for verification.
8. **Withdraw Dose According to Protocol** Draw up the calculated activity of the radiotracer:
  - 3.5 MBq/kg ± 10%, not to exceed 400 MBq
9. **Draw Up Radiotracer** Using either 1 mL, 3 mL, or 5 mL plastic syringe (depending on volume required), with the appropriate syringe shield, draw up the radiotracer dose.
10. **Record Pre-Injection Activity** Measure and record the radioactivity level of the syringe using a dose calibrator, note the exact time, and place the syringe in a lead shielded syringe carrier.
11. **Inject Radiotracer** Slowly inject the radiotracer through the IV line, followed immediately by a 10 mL flush of sterile saline to ensure complete administration.
12. **Secure Radioactive Materials** After injection, disconnect and place the used syringe securely in a lead carrier box.
13. **Remove IV Catheter** Remove the IV catheter carefully using appropriate radiation precautions to prevent contamination.
14. **Record Injection Time** Accurately document the time of injection by checking a calibrated wall clock.
15. **Measure Residual Activity** Place the used syringe back into the dose calibrator to measure residual activity and record the value and time.
16. **Dispose of Waste Safely** Dispose of the syringe and any radioactive waste in a designated radioactive waste container following institutional guidelines.
17. **Uptake Period** Instruct the patient to rest quietly in the ‘hot waiting room’ for 50 minutes to 1 hour to allow for optimal tracer uptake.
18. **Position for Imaging** After the uptake period, escort the patient to the camera room and assist them to lie supine on the PET scanning table.
19. **Final ID Check** Perform a final patient ID check and confirm details against the scanner console before proceeding.
20. **Start PET Scan** Once the correct patient is selected on the console, begin the PET scan according to protocol.
21. **Post-Scan Instructions** After the scan, escort the patient to the exit area and advise they may resume normal eating and drinking.
22. **Radiation Safety Advice** Reiterate radiation safety precautions: Avoid close contact with children and pregnant women for at least 8 hours after the scan.
23. **Encourage Hydration** Advise the patient to hydrate well over the next 24 hours to promote clearance of the radiotracer via urination.
24. **PET Image Reconstruction** Perform PET image reconstruction using Q.Clear.

## 2. Quantification of FDG avid intra-thoracic lymph node using the 3D Slicer platform

In this study, anonymised PET-CT DICOM images were analysed using 3D Slicer version 5.4.0 (https://www.slicer.org/). PET image data was extracted by a consultant thoracic PET-CT radiologist (reader 1) and a trained respiratory registrar (reader 2). PET-CT images with intra-thoracic lymph node FDG uptake above the physiological reference FDG uptake were included for analysis. Physiological reference was determined by liver SUVmean + 3 standard deviations (SD)^10^.

1. **Install Extensions**
  1.1. **Open the Extensions Manager.**
  1.2. Install the following extensions: PETDICOMExtension, PETTumorSegmentation, PETLiverUptakeMeasurement, and PET-IndiC.
  1.3. After installation, restart 3D Slicer.
2. **Upload Anonymised PET-CT DICOM Images to 3D Slicer**
  2.1. Go to Add DICOM Data → Import DICOM files.
3. **Liver Quantification (reference tissue)**
  3.1. **Method I**
    3.1.1. Go to Quantification → PET Liver Uptake measurement.
    3.1.2. In the Input Volume dropdown menu, select PET image.
    3.1.3. In the Output Volume dropdown menu, select “Create New LabelMapVolume.”
    3.1.4. Click “Segment Reference Region”.
    3.1.5. Check the resulting region for correctness. If automated liver segmentation fails, proceed to Method II or III
    3.1.6. Save the mean SUV and standard deviation values under Measurements.
  3.2. **Method II**
    3.2.1. Go to Markups.
    3.2.2. Click “Create new ROI” and create a new ROI around the liver.
    3.2.3. Go to the PET Liver Uptake measurement
    3.2.4. In the Input Volume dropdown menu, select the PET image.
    3.2.5. In the Output Volume dropdown menu, select “Create New LabelMapVolume.”
    3.2.6. In the Region dropdown menu, select the new ROI (default name is R)
    3.2.7. Click “Segment Reference Region.”
    3.2.8. Check the resulting region for correctness.
    3.2.9. Save mean SUV and standard deviation values under Measurement.
  3.3. **Methods III (manual measurement)**
    3.3.1. Go to Segment Editor.
    3.3.2. In the Source volume dropdown menu, select the PET image.
    3.3.3. Click Add.
    3.3.4. Use Paint (select Sphere brush) to create VOI on a homogeneous region of the liver parenchyma.
    3.3.5. Go to Quantification → Segment Statistics.
    3.3.6. In the Scalar volume dropdown menu, select PET image.
    3.3.7. Under Advanced Options, select PET Volume Statistics (or Scalar Volume Statistics) and choose Mean and Standard Deviation.
    3.3.8. Click Apply.
4. **Segmentation of Hot Intrathoracic Lymph Nodes**
  4.1. Create a new segmentation using Segment Editor.
  4.2. Use the Threshold tool. Set the lower limit to SUVmean_liver_ + 3 × SD_liver_. Click Apply. This will highlight all regions in the body above the threshold.
4.3. Exclude areas outside the intrathoracic lymph nodes.
  4.3.1. **Method I: Manual clean up**
  Use the Scissors tool with “Erase Inside” or “Erase Outside” in Segment Editor to remove regions outside the lymph node areas.
  4.3.2. **Method II: “Islands” effect**
  Apply the “Islands” effect in Segment Editor. Use the “Split Islands to Segments” function to divide disconnected hot regions into separate segments. Delete segments outside the lymph node area. Small noise regions may require additional clean up.
5. **Quantification of segmented lymph nodes**
  5.1. Go to Quantification → Segment Statistics.
  5.2. In the Scalar Volume dropdown menu, select the PET image.
  5.3. Under Advanced options, select PET Volume Statistics (or Scalar Volume Statistics) and choose the measurements. Note that TLG need to be calculated manually using scalar volume statistics.
  5.4. Click Apply.

## RESULTS

### Study cohort

Sixteen QFT-Plus positive contacts from nine microbiologically confirmed pulmonary TB index cases (eight smear-positive) had a PET-CT scan. Eleven of sixteen QFT-Plus positive participants had FDG uptake in intrathoracic lymph nodes above the threshold and were included for the analysis. All eleven were contacts of smear-positive pulmonary TB from seven index cases. Median age was 32 (IQR 18 – 46) years. Five (45.5%) were male. Nine (81.8%) were born outside of UK. Seven (63.6%) were never smokers, three (26.3%) were current smokers, and one (9.1%) was an ex-smoker.

Eight participants had repeat PET-CT at three months. Thus, a total of 19 PET-CT scans from eleven treatment naïve contacts were included.

### Inter-platform reliability of clinical (XD3) and research (3D Slicer) platforms

A high agreement (CCC 0.95 – 0.99) was observed between XD3 and 3D Slicer for ITLN SUVmax, MV, and TLG. Moderate-to-high agreement (CCC 0.90 – 0.95) was achieved for the liver SUVmean and ITLN SUVmean. Liver SUVmean + 3 SDs showed fair agreement (CCC = 0.85) (Table 1).

**Table 1.** Agreement between clinical (XD3) and research (3D Slicer) platforms. SUVmax: maximum standardised uptake value, SUVmean: mean standardised uptake value, MV: metabolic volume, TLG: total lesion glycolysis, SD: standard deviation.

|  | Concordance correlation coefficient | 95% confidence interval | Bias correction factor (Cb) |
| --- | --- | --- | --- |
| Liver SUVmean | 0.90 | 0.75-0.96 | 0.97 |
| Liver SUVmean+3SDs | 0.85 | 0.64-0.94 | 1.00 |
| SUVmax | 0.99 | 0.98-1.00 | 0.99 |
| SUVmean | 0.91 | 0.77-0.96 | 0.99 |
| MV | 0.97 | 0.93-0.99 | 1.00 |
| TLG | 0.99 | 0.98-1.00 | 1.00 |

### Inter-operator reliability of 3D Slicer

Reader 1 and Reader 2 demonstrated excellent agreement (ICC > 0.95) across all quantitative metrics extracted using 3D slicer (Table 2).

**Table 2.**
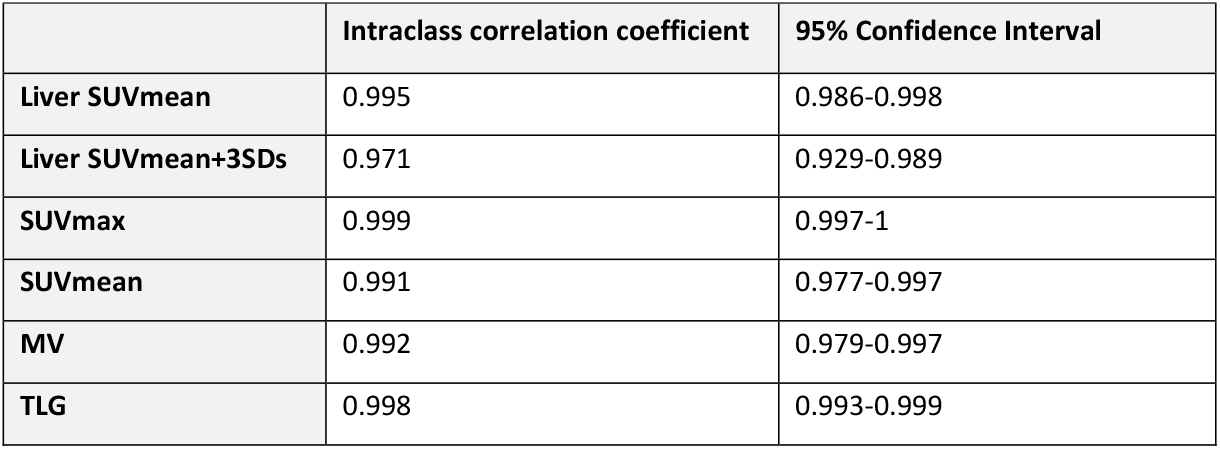
Agreement between Reader 1 and Reader 2 using research platform (3D Slicer). SUVmax: maximum standardised uptake value, SUVmean: mean standardised uptake value, MV: metabolic volume, TLG: total lesion glycolysis, SD: standard deviation.

### Application to longitudinal PET-CT features

Eight untreated QFT-Plus positive participants underwent follow-up PET-CT scans at 3 months. Of these, three individuals received anti-TB treatment. Two commenced on treatment shortly after the follow-up scan: one with confirmed Mtb infection from an intrathoracic lymph node sample, and one who developed a new lung lesion on the 3-month PET scan that resolved with treatment. The third individual developed symptoms of TB during the 2-year prospective follow-up period, with infection confirmed at the site of FDG avidity on the baseline scan.

Two participants that were treated for TB after 3 months demonstrated increased SUVmax, SUVmean, MV and TLG (Figure 1: A1, B1, C1, D1). However, the person who was diagnosed with TB at 2 years showed decreasing SUVmax, SUVmean, and TLG (Figure 1: A1, B1, D1). The MV in this patient remained unchanged (1.50 cm3 to 1.52 cm^3^) (Figure 1: C1). However, physiological FDG uptake measured by Liver SUVmean + 3 SDs was significantly lower at the 3-month scan (3.69 to 2.04), raising the possibility of variability in physiological FDG uptake obscuring interpretation of changes in pathological uptake. Next, we standardised PET parameters to physiological activity to mitigate this variation. Normalising to FDG uptake in the liver revealed increased SUVRmax, and to a lesser extent, increased SUVRmean and normalised TLG in the individual diagnosed with TB at 2 years. (Figure 1: A2, B2, D2). Participants who remained disease-free demonstrated decreasing SUVmax, SUVmean, MV, and TLG between baseline and 3 months (Figure 1: A1, B1, C1, D1).

**Figure 1.**
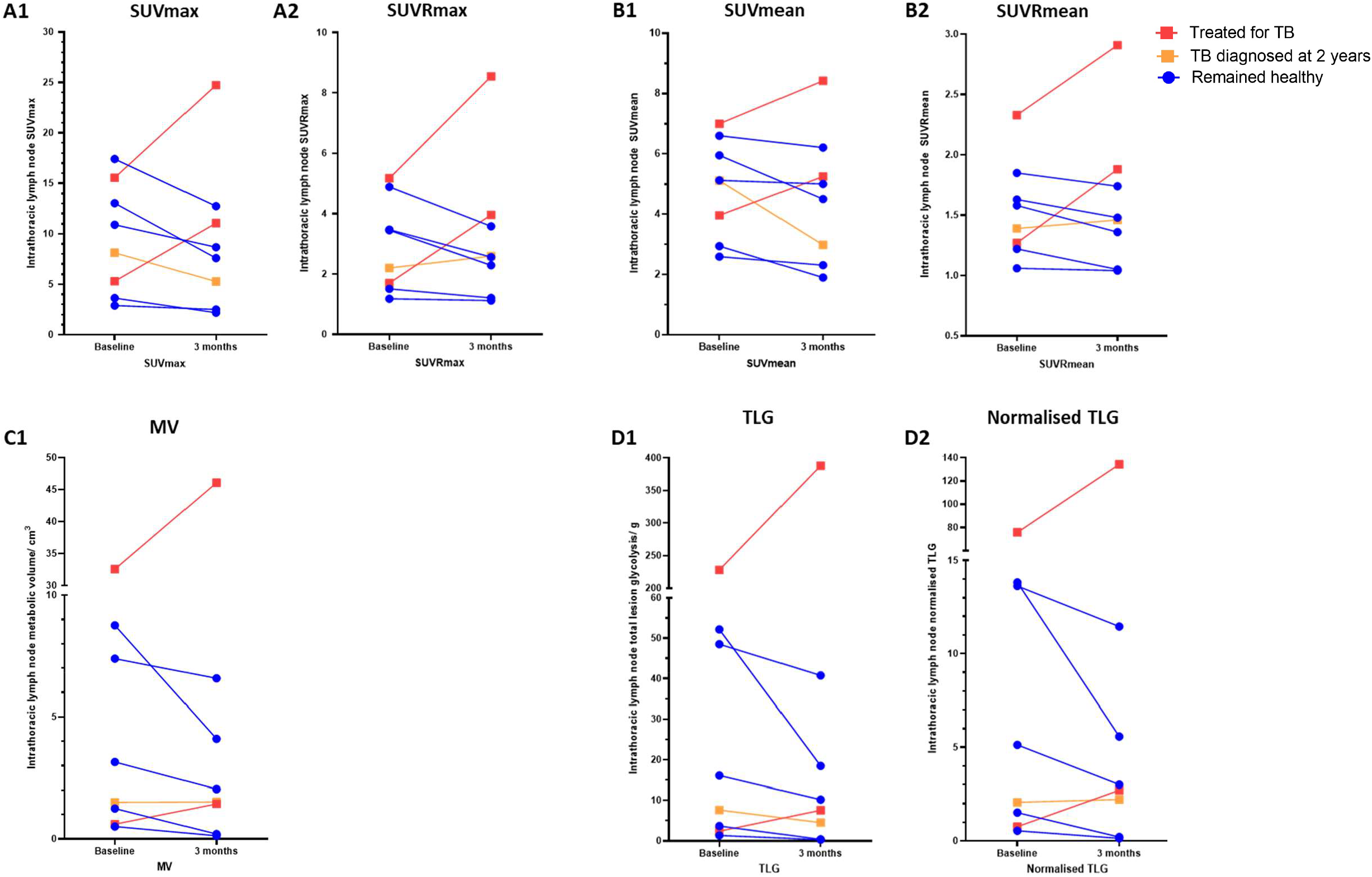
Metabolic activity of intrathoracic lymph nodes at baseline and 3 months, stratified based on clinical outcome. SUVmax: maximum standardised uptake value. SUVRmax: SUV ratio maximum (SUVmax normalised to liver uptake), SUVmean: mean standardised uptake value, SUVRmean: SUV ratio mean, MV: metabolic volume, TLG: total lesion glycolysis

## DISCUSSION

We evaluated semi-automated PET quantification in pulmonary TB contacts using a research imaging platform and observed substantial agreement with clinical software across PET parameters (SUVmax, SUVmean, MV and TLG), and moderate agreement in Liver SUVmean + 3SD threshold. We showed excellent inter-operator agreement in all PET parameters between two independent and blinded readers with a semi-automated approach. Importantly, this was achieved with one reader having no formal specialist radiology training, indicating that automation with 3D Slicer effectively mitigates experiential inter-operator variability.

Normalising PET parameters to a liver threshold appeared to increase the sensitivity of the analysis and revealed an increase in metabolic activity between baseline and 3 months that was otherwise obscured by lower levels of physiological uptake during the second scan. FDG uptake can be influenced by various biological and technical factors, including patient stress, physical activity, blood glucose level, and the interval between FDG injection to scanning^19^. Longitudinal changes on PET-CT in TB contacts are likely less pronounced than those observed during treatment of active TB. Therefore, standardisation using reference tissue and strict adherence to the imaging protocol are essential to improve repeatability and reliability of imaging analyses in TB infection studies^20^. In this context, semi-automated methods and normalisation to a reference organ offer additional value by reducing operator-dependent variability and improving the consistency of quantitative assessments across time points.

An important unknown in the interpretation of PET-CT scans in TB infection is defining clinically meaningful metabolic activity and thresholds for meaningful change over time. In our study, all PET parameters decreased at three months among individuals who remained healthy. However, baseline quantitative values overlapped between those who remained healthy and those who received treatment. This suggests that the trend over time may be more important than measurement at a single time point. Future studies should evaluate multiple quantitative measures of PET-CT metabolic activity over time and correlate these with pre-defined clinical measures to determine the optimal parameters and establish reference thresholds that facilitate standardised interpretation.

We focused on the quantification of intrathoracic lymph nodes due to a limited number of lung parenchymal abnormalities. Findings from our study showed that early TB infection is predominantly confined to intrathoracic lymph nodes, which is consistent with a study conducted in the USA^6^. In our cohort, only four participants demonstrated lung parenchymal FDG uptake at baseline, with low metabolic activity (mean SUVmax = 2.7), and the lesions were too small for reliable volumetric analysis and characterisation. In addition, only one out of eight QFT-Plus positive participants who underwent serial scans showed lung parenchymal FDG uptake at baseline. In contrast, a study conducted in a high TB burden setting (South Africa) reported lung parenchymal abnormalities in approximately half of the household TB contacts, while FDG-avid intrathoracic lymph nodes were observed in 21.2%, and 9.3% of those with normal lungs^5^. The reason for the difference observed in PET-CT features between our cohort (low TB burden) and the cohort from the high TB burden area is unclear. One possibility is the high prevalence of past TB infection in the South African cohort, evidenced by a high proportion of QFT-Plus positivity (82%). A non-human primate (NHP) study comparing Mtb reinfection and infection in Mtb-naïve NHPs showed that primary infection provides protection against reinfection, including the prevention of dissemination to intrathoracic lymph nodes^21^. Other potential contributing factors include differences in Mtb strains, host genetic factors, prevalence of smoking in study cohorts, and environmental factors. Future TB infection studies employing PET-CT should consider Mtb and cohort characteristics and develop approaches to integrate changes in lung parenchyma and intrathoracic lymph nodes.

In summary, semi-automated PET quantification can offer a valuable approach for assessing metabolic changes in TB infection with high inter-operator reliability. Normalisation to reference tissues enhances analytical sensitivity in detecting early metabolic progression. Increasing metabolic activity in early infection, when corrected to a reference organ, is a promising indicator of progressive TB infection.

## Data Availability

Requests for additional data not included in the paper will be considered upon request but may be subject to additional ethical review.

## ACKNOWLEDGMENTS

This is a summary of independent research funded by the University of Leicester MRC Confidence in Concept award to P.H. and carried out at the National Institute for Health and Care Research (NIHR) Leicester Biomedical Research Centre (BRC). The views expressed are those of the author(s) and not necessarily those of the funders, NHS, the NIHR or the Department of Health and Social Care.

## DISCLOSURES

All authors report no potential conflicts to declare.

## Contributors

P.H. and J.W.K. conceived the study. A.K. and J.W.K. developed the protocol and extracted data from PET-CT scans. P.H., J.K., J.W.K., and I.N. contributed to the enrolment of participants and clinical data collection. J.W.K., A.K., and P.H. have accessed and verified the data. J.W.K. wrote the manuscript with contributions from all the authors. All authors had full access to all the data in the study and take final responsibility for the decision to submit for publication.

## Notes

### Competing Interest Statement

The authors have declared no competing interest.

### Funding Statement

The study was funded by University of Leicester MRC Confidence in Concept award to P.H..

### Author Declarations

The study was approved by the Research Ethics Committee for East Midlands, Nottingham 1, Nottingham, UK (REC 15/EM/0109).

